# Local public health officials and COVID-19: Evidence from China

**DOI:** 10.1101/2020.07.18.20156828

**Authors:** John (Xuefeng) Jiang, Maobin Wang

**Author notes:** Contribution: JJ* and MW both planned the study. MW carried out the analyses. JJ wrote up the results and is responsible for the overall content as guarantor. Competing interests: Dr. Jiang and Dr. Wang have nothing to disclose. Funding:* The current study was not funded by any agencies. Ethical approval is not required*. Patient and Public Involvement*: No patients were involved in this study.

## Abstract

Local public health authorities are at the forefront of fighting COVID-19. They monitor its spread in the local population and advise the local government on whether to close schools and businesses. Examining their role in battling COVID-19 will inform the public how best to prepare for a public health emergency. This study examined whether more competent local public health officials in China are more effective in fighting COVID-19, where competence was measured by the public health officials’ professional background. Only 38% of the heads of the public health departments of Chinese cities have a medical background. Cities with medical professionals as the head of public health departments had lower infection rates and death rates from COVID-19. The results were significant only at the start of the outbreak. Our results suggest that we should staff local public health authorities with competent professionals to better combat a pandemic.

## Introduction

The spread of COVID-19 reveals that most countries are ill-prepared for a severe public health emergency. It is critical to understand how the public health sectors can help to combat a pandemic like COVID-19. In a new book, *The Premonition*, Michael Lewis recounted the story of Charity Dean, the Public Health Officer of Santa Barbara County, who has successfully stopped the spread of many infectious diseases. Large sample evidence of the impact of public health officials is still rare. This study examined whether more competent local public health officials lead to lower infection rates and deaths rate of COVID-19, using data from China.

The Chinese setting offers an imperfect but relatively objective measure of the competence of the local health officials. In developed countries, public health officials almost always have professional credentials. However, this is not typically the case in China. For a long time, the Chinese government has focused on fostering economic growth. Government posts that are not directly related to economic growth are not viewed as important or prestigious. As a consequence, many local public health offices were not headed by people with medical or public health background.

We collected the professional background of the directors of the public health departments of 350 Chinese cities, which include 87% of the Chinese population. Excluding Wuhan, the epicenter of COVID-19, we analyzed the infection rates and death rates from COVID-19 between 131 Chinese cities whose public health departments are led by medical professionals and 218 cities whose public health departments are led by nonprofessionals. We employed a multivariate regression controlling for the number of people that traveled from Wuhan to each city, the local economic development, and the number of hospital beds.

Only 38% of the heads of the public health departments of Chinese cities have a medical background. Chinese cities whose public health departments are led by medical professionals had 21 fewer confirmed cases per 10 million as of January 31, 2020 [95% CI, -40 to -3], 58 fewer cases per 10 million in the next ten days [95% CI, -116 to 0], similar new cases between February 11 and February 20, 2020, and 3 fewer deaths per 10 million as of February 20, 2020 [95% CI, -7 to 0].

We find that Chinese cities whose public health departments are headed by medical professionals were associated with lower infection rates and fewer death rates from COVID-19. The results were significant only at the start of the outbreak. Our results suggest that we should staff local public health authorities with competent professionals to better combat a pandemic.

## Main Text

On the one hand, it seems unambiguous that more competent public health officials should lead to better outcomes. On the other hand, public health officials report to politicians who might ignore their advice due to other considerations. It is challenging to link the outcome to the quality of local public health officials. For example, in the United States, researchers found that state governors’ party affiliation was the most important determinant of when states adopted social distancing policies.^*1*^ In China, the failure to sound the alarm early about COVID-19 was attributed to local officials’ “political aversion to sharing bad news”.^2^ However, once President Xi publicly called for local governments to contain the outbreak, public health officials and politicians’ incentives were aligned. Because most Chinese cities had no experience in dealing with infectious diseases, the local public health officials played a critical role in shaping their jurisdiction’s early responses. Their influences can be seen by the accountability actions where some local public health officials were demoted while others were promoted for their performance in the pandemic. These news items suggest that the early stage of the outbreak in China was a powerful setting to assess the importance of local public health officials.

It is also difficult to measure the competence of local public health officials. The China setting offers us an imperfect but relatively objective measure. In developed countries, public health officials almost always have professional credentials. However, this is not typically the case in China. For a long time, the Chinese government has focused on fostering economic growth. Government posts that are not directly related to economic growth are not viewed as important or prestigious. As a consequence, many local public health offices were not headed by people with medical or public health backgrounds.

In addition, a recent government reshuffle added more bureaucrats to the public health offices. In 2013, following the merger of the Ministry of Health and the National Population and Family Planning Commission at the national level, local public health offices also combined with local family planning commissions, which were created to enforce the one-child policy in the 1980s. The merger led many former bureaucrats from the family planning commissions to head the combined public health departments, potentially diluting their mission in disease prevention. We use public health officials’ professional backgrounds to proxy for competence. Though this measure is imperfect, it has anecdotal supports. For example, the head of the public health authority of Huangguang City, which had the second most confirmed cases in Hubei province, was dismissed for causing problems such as “insufficient screening for suspected cases, slow progress of tests and lack of testing personnel.”^3^ The media noted that she studied law in college and had no working experience in healthcare.

### Unit of analyses

In China, prefectures are administrative divisions that are smaller than provinces and bigger than counties. The epicenter of the COVID-19 outbreak, Wuhan, is a prefecture in Hubei province with more than 11 million residents. We chose prefectures as the unit of analyses because the COVID-19 cases were consistently reported by prefectures. Their public health departments carried out the tests of the virus, communicated with the local community about confirmed cases, and allocated medical personnel across different hospitals in the area. The preventive measures, such as discouraging public gatherings and closing restaurants and schools, were also announced by the prefecture governments. Since most prefectures are middle-sized cities, we refer to cities and prefectures interchangeably in the rest of this article.

### Local public health chiefs with medical backgrounds

We collected the resumes of directors of the public health department from the individual city government’s website. China has 31 provinces. We excluded Xizang (Tibet) due to its special governance status and lack of information disclosed on its government website. Among 412 cities of the remaining 30 provinces in China, we successfully identified the detailed background of the chief public health officials of 350 cities, which included 87% of the Chinese population. We coded public health officials as medical professionals if they had a medical degree or started their career in a hospital, epidemic prevention center, or other healthcare-related fields. Among the 350 public health chiefs, only 38% were medical professionals. Figure 1 reported the percentage of public health chiefs who were medical professionals and the average GDP per capita for each province. The graph indicated that affluent areas were more likely to appoint medical professionals as public health chiefs. The Pearson correlation was about 0.6 for these two variables. We plotted a linear regression line in the graph in Figure 1 (the underlying data were reported in the appendix). Beijing and Guangdong, two places that suffered the most from the 2003 SARS outbreak, had a higher than expected proportion of medical professionals leading local public health departments. After experiencing a public health crisis, perhaps the local governments learned to appreciate the importance of qualified public health officials.

**Fig. 1.**
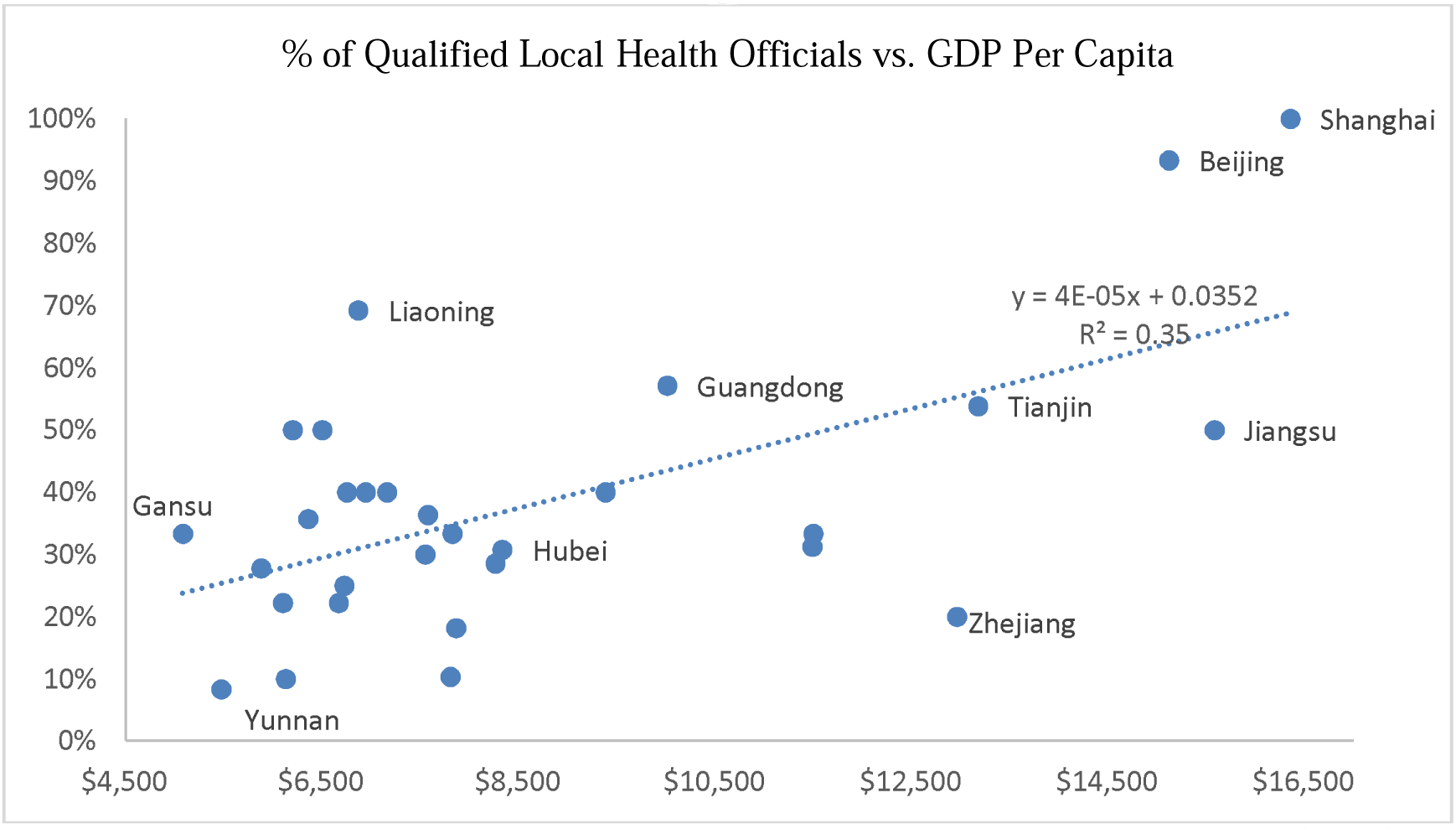

### Infection rates and death rates from COVID-19

We excluded Wuhan from the following analysis because of its extreme situation.^4^ As of February 20, 2020, the confirmed cases in Wuhan were 70 percent greater than those in the rest of our sample areas combined. The deaths from COVID-19 in Wuhan were more than three times greater than those of the rest of our sample areas. Including Wuhan would distort this study’s inference. Also, only the deaths from Wuhan have been revised recently, which indicated that the data collection in Wuhan might be incomplete at the start of the pandemic. Excluding Wuhan reduced the measurement errors in these figures.

We analyzed the infection rate over time to assess the impact of local public health officials. As the number of confirmed cases climbed, all local governments mimicked each other and adopted the same policy. If competent local public health officials had any impact, more likely it would be observed in the early period of the COVID-19 outbreak.

The spread of COVID-19 was highly associated with the number of people who migrated from Wuhan.^*5*^ Accordingly, we controlled for the total number of people who traveled from Wuhan to each city between January 1 and January 22, 2020. The data were provided by Baidu, one of the largest internet companies in China, which reported aggregate daily migration data among cities based on users’ map applications. Baidu made the data from January 1 2020 to May 8 2020 publicly available to facilitate research on COVID-19. The data are archived at Harvard Dataverse under the name Baidu Mobility Data.

Wuhan is the capital city of Hubei province. To capture the close connection among prefectures in Hubei, we included a Hubei indicator. Since affluent cities were more likely to appoint former medical practitioners to head their public health departments, we included a city’s GDP per capita to isolate the impact of professional public health officials from an area’s economic development. We also construct a city efficiency measure based on Tang et al. (2014)^6^. More efficient local governments are associated with faster urban growth and more exports (Reinecke and Schmerer, 2017; Rodriguez-Rose and Zhang 2019)^7,8^. More efficient local government might be better at dealing with a health emergency. When analyzing the death rates from COVID-19, we also included the number of beds per thousand to proxy for a city’s medical resources. In sum, here are the models we use. We estimate them by the ordinary least squares using STATA 16.

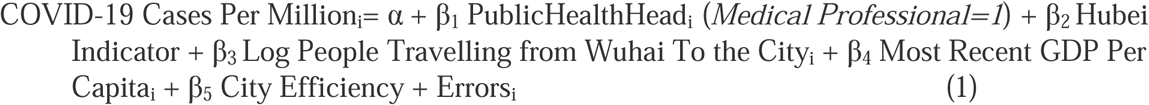

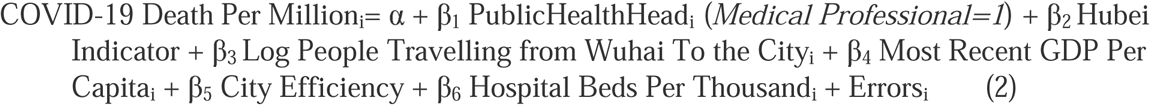

### Descriptive statistics

Table 1 reported the descriptive statistics of all variables in our analyses. The table note includes all the data sources. As of January 31, 2020, the average infected cases per 10 million in each city other than Wuhan were 54, with 6426 cases in total. New cases increased to 122 per 10 million in the ten days ending on February 10, with 14,845 cases in total. In the next ten days, only 55 new cases per 10 million were reported, with 5,558 cases in total. As of February 20, 2020, there were four deaths per 10 million resulting from COVID-19 (464 deaths in total). From January 1 to January 22 of 2020, Baidu recorded that people traveled from Wuhan to other cities of China more than 91 million times. We took the natural logarithm of this variable to deal with the skewness. The average GDP per capita was $8,900 at the end of 2017, the latest number available. On average, there were 5 hospital beds per thousand people in each city.

**Table 1.** Descriptive statistics for 349 prefectures (excluding Wuhan)

| Variables | mean | median | std | p25 | p75 |
| --- | --- | --- | --- | --- | --- |
| COVID-19 cases per 10 million as of 01/31/2020 | 54 | 18 | 155 | 7 | 41 |
| New cases per 10 million 02/01/2020 -02/10/2020 | 122 | 30 | 470 | 12 | 69 |
| New cases per 10 million 02/11/2020 -02/20/2020 | 55 | 8 | 337 | 0 | 21 |
| COVID-19 deaths per 10 million as of 02/20/2020 | 4 | 0 | 25 | 0 | 0 |
| Public health dept. headed by medical professional (Yes=1) | 0.38 | 0.00 | 0.48 | 0.00 | 1.00 |
| Located in Hubei Province (Yes=1) | 0.03 | 0.00 | 0.18 | 0.00 | 0.00 |
| Natural log of people (000) traveling fr. Wuhan 01/01-01/22 | 4.07 | 4.12 | 1.32 | 3.12 | 4.87 |
| GDP per capita (000 \$) | 8.90 | 7.20 | 5.74 | 5.05 | 10.80 |
| City efficiency (from 1 to 4, 4 is the highest efficiency) | 1.45 | 1.00 | 0.86 | 1.00 | 2.00 |
| Hospital beds per thousand people | 5.2 | 5.0 | 1.7 | 4.2 | 5.7 |

Note: confirmed cases and deaths of COVID-19 from each city were gathered from dingxiangyuan (ncov.dxy.cn), a professional healthcare website whose primary resource is newspapers and national and local governments. The city population and Gross Domestic Product (GDP) were from the 2018 China Statistical Yearbook. The number of times that people traveled from Wuhan to each city from January 1, 2020 to January 22, 2020 (before Wuhan was quarantined) was reported by Baidu, a leading internet company in China, which aggregated users' location data on a daily basis. Following the 2019 Research Report on Local Government Efficiency in China, we code a city's efficiency as 4 if it is ranked from 1 to 20; 3 if ranked from 21 to 50; 2 if ranked from 51 to 100; 1 if not ranked. The ranking is based on a city government's performance in science, education, and health. The number of hospital beds was from CEIC Data, a data provider of Chinese statistics.

### Main Results

We used multivariate regressions to explain the confirmed cases and deaths from COVID-19. The analyses were carried out in STATA version 14. As shown in Table 2, professional public health chiefs were associated with 21 fewer confirmed cases of COVID-19 as of January 31, 2020 (two-tailed p-value=0. 02), and 58 fewer new cases between February 1 and February 10, 2020 (two-tailed p-value=0. 05). For the next 10 days, professional public health chiefs were not significantly associated with fewer new COVID-19 cases. The results were consistent with the hypothesis that the impact of professional public health chiefs was more critical in the early days of the pandemic episode.

**Table 2.** Multivariate regression to explain the COVID-19 infection rates and death rates among 349 Chinese cities

| VARIABLES | (1)<br>COVID-19<br>cases per 10 mi<br>as of<br>01/31/2020 | (2)<br>New cases per 10 mi<br>btw 02/01 and 02/10 | (3)<br>New cases per 10 mi<br>btw 02/11 and 02/20 | (4)<br>Deaths per<br>10 mi as of<br>02/20 |
| --- | --- | --- | --- | --- |
| Public health<br>head is medical<br>professional | -21.4**<br>(0.022) | -58.2**<br>(0.048) | -35.7<br>(0.146) | -3.3*<br>(0.080) |
| Located in Hubei | 613***<br>(0.000) | 1,954***<br>(0.000) | 1,038***<br>(0.010) | 99***<br>(0.000) |
| Ln of people<br>travelling from<br>Wuhan | 20.9***<br>(0.000) | 39.1***<br>(0.000) | 16.1**<br>(0.011) | 1.0**<br>(0.024) |
| GDP per capita<br>(000 \$) | 1.1<br>(0.126) | 2.4<br>(0.196) | 1.9<br>(0.370) | 0.2<br>(0.249) |
| City_efficiency | -1.1<br>(0.759) | -5.5<br>(0.622) | 0.3<br>(0.973) | -0.3<br>(0.690) |
| Hospital beds per<br>thousand |  |  |  | -0.7*<br>(0.060) |
| Constant | -53***<br>(0.000) | -95.3***<br>(0.000) | -50.4<br>(0.228) | -0.3<br>(0.953) |
| Observations | 349 | 349 | 349 | 349 |
| Adj R-squared | 0.73 | 0.70 | 0.36 | 0.55 |

Robust p-value in parentheses; All variables are defined in Table 1
\*\*\* $p \leq 0.01$ , \*\* $p \leq 0.05$ , \* $p \leq 0.10$

As of February 20, 2020, cities whose public health chiefs had medical backgrounds experienced three fewer deaths per 10 million people from COVID-19 (two-tailed p-value=0.08). Figure 2 presents these estimates more vividly. The numbers were generated by STATA’s margins command, which estimated the infected cases and deaths for the two groups of cities holding all other variables at the mean value. We didn’t graph the estimates from February 11 to February 20, 2020, since the difference between the two groups was not statistically significant.

**Fig. 2.**
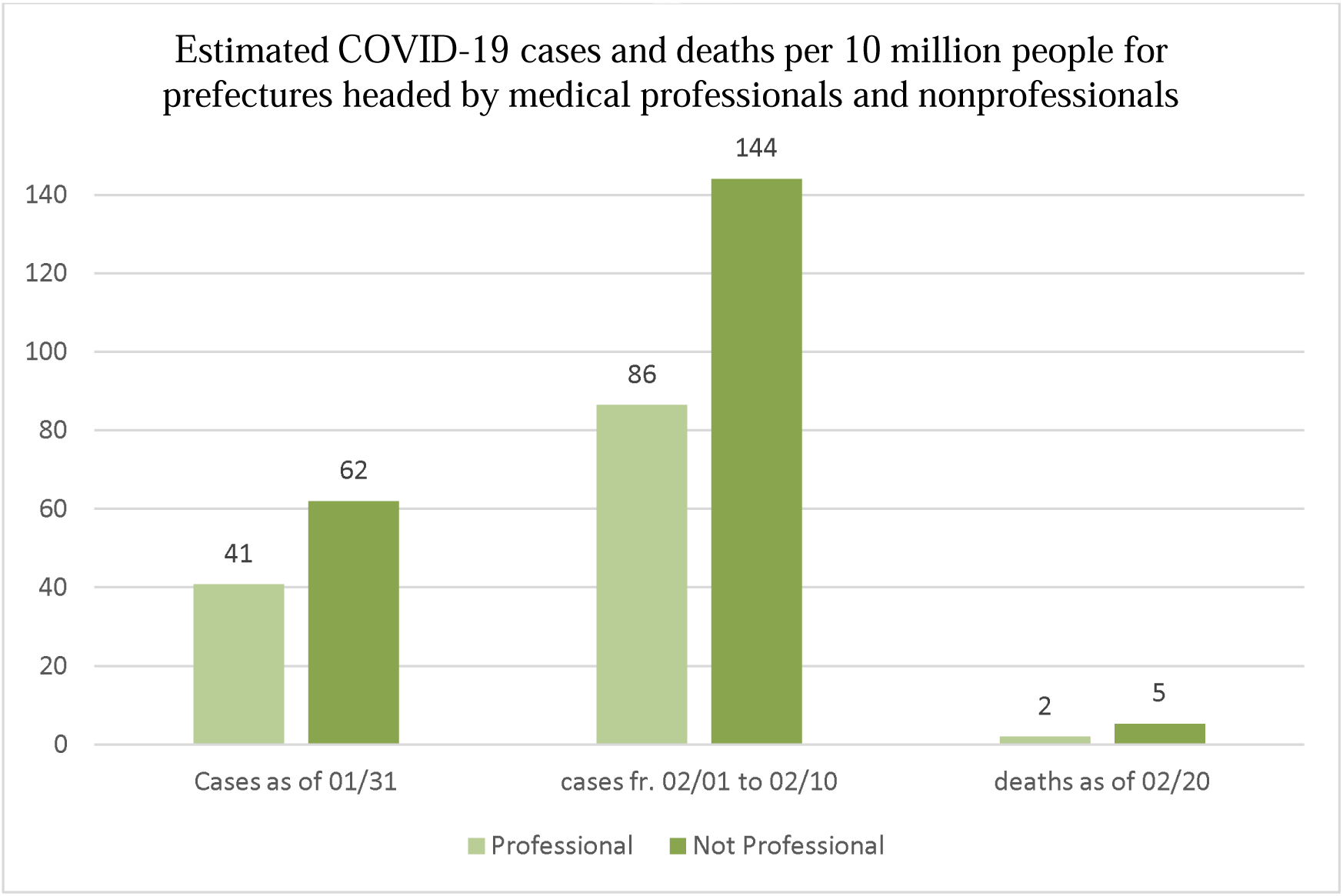

The results also indicated that the Hubei indicator and the number of times people traveled from Wuhan were positively associated with the confirmed cases and the deaths from COVID-19. The number of hospital beds was negatively associated with the deaths from COVID-19, indicating areas with more medical resources could better handle severe cases. The adjusted R^2^, which captured the overall model fit, declined from 0.73 to 0.70 and 0.36 in explaining the infection rates over time. The change indicated that over time the spread of COVID-19 was less associated with imported cases from Wuhan.

### Adding province fixed effects

If cities have different cultures such as individualism vs collectivism, individuals might respond to the same public policies differently. In the United States, people respond differently to the governments’ mask policy and the vaccination requirement. To appropriately measure the effect of local public health officials, it might be important to control for the local culture. We do not know how to measure a city’s local culture. However, the media and the public tend to talk about a group’s personality based on geographic areas. For example, people from northeast China are viewed as having better humor. People from Hubei province are viewed as intelligent and capable. We suspect that cities within one province might be more similar culture-wise than cities between provinces. We add the province fixed effects in the following analyses. In this model specification, cities within the same province are compared with each other. To the extent that the local culture is similar within a province, our approach will be able to eliminate its influence on the local public health officials. We report this set of results in Table 3. The inferences on our variable of interests remain the same.

**Table 3.** Adding Province Fixed Effect Multivariate regression to explain the COVID-19 infection rates and death rates among 349 Chinese cities

| VARIABLES | (1)<br>COVID-19<br>cases per 10 mi<br>as of<br>01/31/2020 | (2)<br>New cases per 10 mi<br>btw 02/01 and 02/10 | (3)<br>New cases per 10 mi<br>btw 02/11 and 02/20 | (4)<br>Deaths per<br>10 mi as of<br>02/20 |
| --- | --- | --- | --- | --- |
| Public health<br>head is medical<br>professional | -17.9*<br>(0.090) | -66.4*<br>(0.056) | -38.9<br>(0.164) | -3.6*<br>(0.089) |
| Located in Hubei | 580.7***<br>(0.000) | 1761.4***<br>(0.000) | 909.6**<br>(0.014) | 88.1***<br>(0.001) |
| Ln of people<br>travelling from<br>Wuhan | 34.4***<br>(0.000) | 92.0***<br>(0.005) | 54.7*<br>(0.073) | 4.4**<br>(0.040) |
| GDP per capita<br>(000 \$) | 1.6*<br>(0.056) | 2.9<br>(0.197) | 2.3<br>(0.373) | 0.2<br>(0.270) |
| City_efficiency | -2.6<br>(0.534) | -8.8<br>(0.535) | -0.3<br>(0.975) | -0.3<br>(0.772) |
| Hospital beds per<br>thousand |  |  |  | -0.9<br>(0.104) |
| Constant | -138.2***<br>(0.005) | -351.4**<br>(0.022) | -253.2<br>(0.138) | -16.7*<br>(0.072) |
| 29 Province Indicator Variables Added |  |  |  |  |
| Observations | 349 | 349 | 349 | 349 |
| Adj R-squared | 0.713 | 0.679 | 0.312 | 0.520 |

Robust p-value in parentheses; All variables are defined in Table 1
\*\*\* $p \leq 0.01$ , \*\* $p \leq 0.05$ , \* $p \leq 0.10$

## Limitations and Discussions

We found that Chinese cities whose public health chiefs had medical backgrounds were associated with lower infections and fewer deaths from COVID-19 at the early period of the outbreak. Our evidence suggested that public health officials’ quality was important for local governments to combat a public health emergency.

Our measure of competent health officials was unambiguous and straightforward, but it was imperfect. Officials with relevant medical backgrounds could be incompetent, while people without the credentials could be effective leaders. In addition, it was possible that the COVID-19 data were not accurately reported in China. Data from Wuhan were more suspicious because of insufficient tests and the chaos caused by a sudden lockdown. We excluded Wuhan from our analyses to reduce the possible measurement errors. The aggregate underreporting of China’s data wouldn’t affect our results because we compared data from different cities in China. A recent study also failed to find any evidence that the COVID-19 data from China were manipulated.^9^ While we have controlled for the main factors that explain why some Chinese cities appointed people with medical backgrounds to head the local public health department, it is still possible that some omitted variables explain our inference.

Though our evidence is based on China, public health officials’ importance is not limited to one country. For example, in western countries, public health officials have more independence from elected officials. They can provide scientific but unpopular opinions. The local public health officials in western countries likely play a more prominent role than their counterparts in China in handling a public health emergency.

We collected the resumes of the directors of public health departments of 350 prefectures (cities) from 30 provinces in China, which include 87% of the Chinese population. For each province, we calculated the percentage of directors with a medical background and the average GDP per capita.

The estimates were made based on the multivariate regression models in Table 2. They were generated by STATA’s margins command. All other variables were taken at their average value.

## Data Availability

Data are from public sources. Detailed summary is in the appendix.

## Acknowledgement

Dr. Jiang thanks the support of the Plante Moran Fellowship, Michigan State University.

---

**Appendix:** Data for Figure 1

| Province Name | Average GDP per capita (\$) | % of Qualified Local Health Officials |
| --- | --- | --- |
| Shanghai | \$ 16,352 | 100% |
| Jiangsu | \$ 15,579 | 50% |
| Beijing | \$ 15,116 | 93% |
| Tianjin | \$ 13,173 | 54% |
| Zhejiang | \$ 12,958 | 20% |
| Fujian | \$ 11,495 | 33% |
| Shandong | \$ 11,485 | 31% |
| Guangdong | \$ 10,008 | 57% |
| Inner Mongolia | \$ 9,380 | 40% |
| Hubei | \$ 8,329 | 31% |
| Xinjiang | \$ 8,259 | 29% |
| Hunan | \$ 7,859 | 18% |
| Hainan | \$ 7,821 | 33% |
| Chongqing | \$ 7,802 | 10% |
| Jiangxi | \$ 7,571 | 36% |
| Shaanxi | \$ 7,545 | 30% |
| Henan | \$ 7,155 | 40% |
| Jilin | \$ 6,938 | 40% |
| Liaoning | \$ 6,862 | 69% |
| Qinghai | \$ 6,746 | 40% |
| Anhui | \$ 6,721 | 25% |
| Hebei | \$ 6,664 | 22% |
| Guizhou | \$ 6,497 | 50% |
| Guangxi | \$ 6,353 | 36% |
| Ningxia | \$ 6,196 | 50% |
| Shanxi (山西) | \$ 6,125 | 10% |
| Heilongjiang | \$ 6,096 | 22% |
| Sichuan | \$ 5,875 | 28% |
| Yunnan | \$ 5,469 | 8% |
| Gansu | \$ 5,078 | 33% |

The average GDP is based on the mean GDP of all the cities in a province from the 2018 China Statistical Yearbook. The % of Qualified Local Health Officials is the proportion of local public health officials that have a medical background based on their resume posted online.

## Notes

Authors declare no potential conflicts of interest with respect to the research, authorship, and/or publication of this article. We have not published any papers from the same study.

### Competing Interest Statement

The authors have declared no competing interest.

### Funding Statement

No external funding.

### Author Declarations

Public available data. Exampt from IRB.

### Summary of Updates

1. Adding measures that control for the local government's efficiency. 2. Adding province-fixed effects to control for any systematic variations within the same province, such as local culture.

## References

1 https://pubmed.ncbi.nlm.nih.gov/32955556/

2 https://www.nytimes.com/2020/03/29/world/asia/coronavirus-china.html

3 https://www.businessinsider.com/analysis-china-hubei-officials-sacked-xi-jinping-protected-2020-2.

4 The dire situation in Wuhan can be traced to the local government’s incompetency. https://www.wsj.com/articles/china-contends-with-questions-over-response-to-viral-outbreak-11579825832.

5 Qiu, Y., X. Chen., and W. Shi. 2020. Impacts of social and economic factors on the transmission of coronavirus disease 2019 (COVID-19) in China. Journal of Population Economics. doi.org/10.1007/s00148-020-00778-2.

6 Tang, R., Tang, T., and Lee, Z. 2014. The efficiency of provincial governments in China from 2001 to 2010: measurement and analysis. Journal of Public Affairs, 14: 142–153. DOI: 10.1002/pa.1518.

7 Reinecke, A. and Schmerer, H. 2017. Government efficiency and exports in China, Journal of Chinese Economic and Business Studies, 15: 249–268. DOI: 10.1080/14765284.2017.1356593.

8 Rodríguez-Pose, A., and Zhang, M. 2019. Government institutions and the dynamics of urban growth in China. Journal of Regional Science. 59: 633–668. DOI: 10.1111/jors.12435

9 https://doi.org/10.1016/j.econlet.2020.109573

